# Pre-vaccination and early B cell signatures predict antibody response to SARS-CoV-2 mRNA vaccine

**DOI:** 10.1101/2021.07.06.21259528

**Authors:** Lela Kardava, Nicholas Rachmaninoff, William W. Lau, Clarisa M. Buckner, Krittin Trihemasava, Felipe Lopes de Assis, Wei Wang, Xiaozhen Zhang, Yimeng Wang, Chi-I Chiang, Sandeep Narpala, Robert Reger, Genevieve E. McCormack, Catherine A. Seamon, Richard W. Childs, Anthony F. Suffredini, Jeffrey R. Strich, Daniel S. Chertow, Richard T. Davey, Michael C. Sneller, Sarah O’Connell, Yuxing Li, Adrian McDermott, Tae-Wook Chun, Anthony S. Fauci, John S. Tsang, Susan Moir

## Abstract

SARS-CoV-2 mRNA vaccines are highly effective, although weak antibody responses are seen in some individuals with correlates of immunity that remain poorly understood. Here we longitudinally dissected antibody, plasmablast, and memory B cell (MBC) responses to the two-dose Moderna mRNA vaccine in SARS-CoV-2-uninfected adults. Robust, coordinated IgA and IgG antibody responses were preceded by bursts of spike-specific plasmablasts after both doses, but earlier and more intensely after dose two. Distinct antigen-specific MBC populations also emerged post-vaccination with varying kinetics. We identified antigen non-specific pre-vaccination MBC and post-vaccination plasmablasts after dose one and their spike-specific counterparts early after dose two that correlated with subsequent antibody levels. These baseline and response signatures can thus provide early indicators of serological efficacy and explain response variability in the population.

---

The pandemic caused by severe acute respiratory syndrome coronavirus 2 (SARS-CoV-2) instigated rapid worldwide COVID-19 vaccine prioritization strategies. Several vaccine candidates were developed, including two vaccines based on novel mRNA platforms (Moderna mRNA-1273 and the Pfizer/BioNTech BNT162b2). Both mRNA vaccines encode a stabilized ectodomain of the spike protein trimer (S-2P) derived from the Wuhan Hu-1 isolate^1^, and are given in two vaccine doses, referred to henceforth as v1 and v2. Both mRNA vaccines have been shown to be highly protective and elicit strong B cell and antibody responses^2,3^, although poorer responses have also been seen in some individuals, such as the elderly^4^ and transplant recipients^5- 7^, raising the question of what determines antibody response levels and whether early correlates of immunity can be defined. Studies on other vaccines have shown that pre-vaccination signatures and early circulating B cell responses involving plasmablasts (PB) and activated memory B cells (MBC) can predict the magnitude and longevity of neutralizing antibodies following vaccination^8-11^. The Pfizer vaccine has been shown to induce robust PB and MBC responses in blood and draining lymph nodes^4,12^, but the extent by which these responses differ across individuals and whether they are associated with antibody levels have not been assessed.

To address gaps in correlates of humoral immunity to mRNA vaccines, we evaluated antibody and B cell responses following vaccination with mRNA-1273 in 21 healthy SARS-CoV-2-uninfected adults (Extended Data Table 1). Blood was drawn serially over a period of ∼60 days (D) and paired serum and cellular assays were performed at each timepoint (Fig. 1a and Extended Data Table 1). Given the fragility of PB, cellular assays were performed on freshly isolated cells while sera were cryopreserved for antibody assays. Antibody binding to S-2P, its receptor-binding domain (RBD) and the nucleoprotein (NP), was measured using a multiplex platform^1^. Strong IgG and IgA responses were induced, starting around D10, to both S-2P and RBD (Fig. 1b), although the magnitude was highly variable across vaccinees at v2D28 (c.v. > 100%), spanning 2-3 orders of magnitude for both IgA and IgG titers (Fig. 1b, right panels). The IgM response was weak across all vaccinees (Fig. 1b). This is consistent with recent reports for the mRNA vaccines^13,14^, yet in contrast to strong responses in patients who recovered from mild to severe COVID-19^14-17^ (Fig. 1c). NP antibodies were also low in vaccinees (Fig. 1c), as expected for SARS-CoV-2-uninfected people. Strong correlations were observed among RBD and S-2P antibodies (Fig. 1d), but the correlation between IgA RBD and IgA S-2P was higher than that between their IgG counterparts. The inhibition of RBD binding to the spike protein receptor ACE2 by serum antibodies, a surrogate for neutralization capacity, also revealed a range of responses (Fig. 1e) that correlated with RBD IgG and IgA binding antibodies (Extended Data Fig. 1a).

**Fig. 1.**
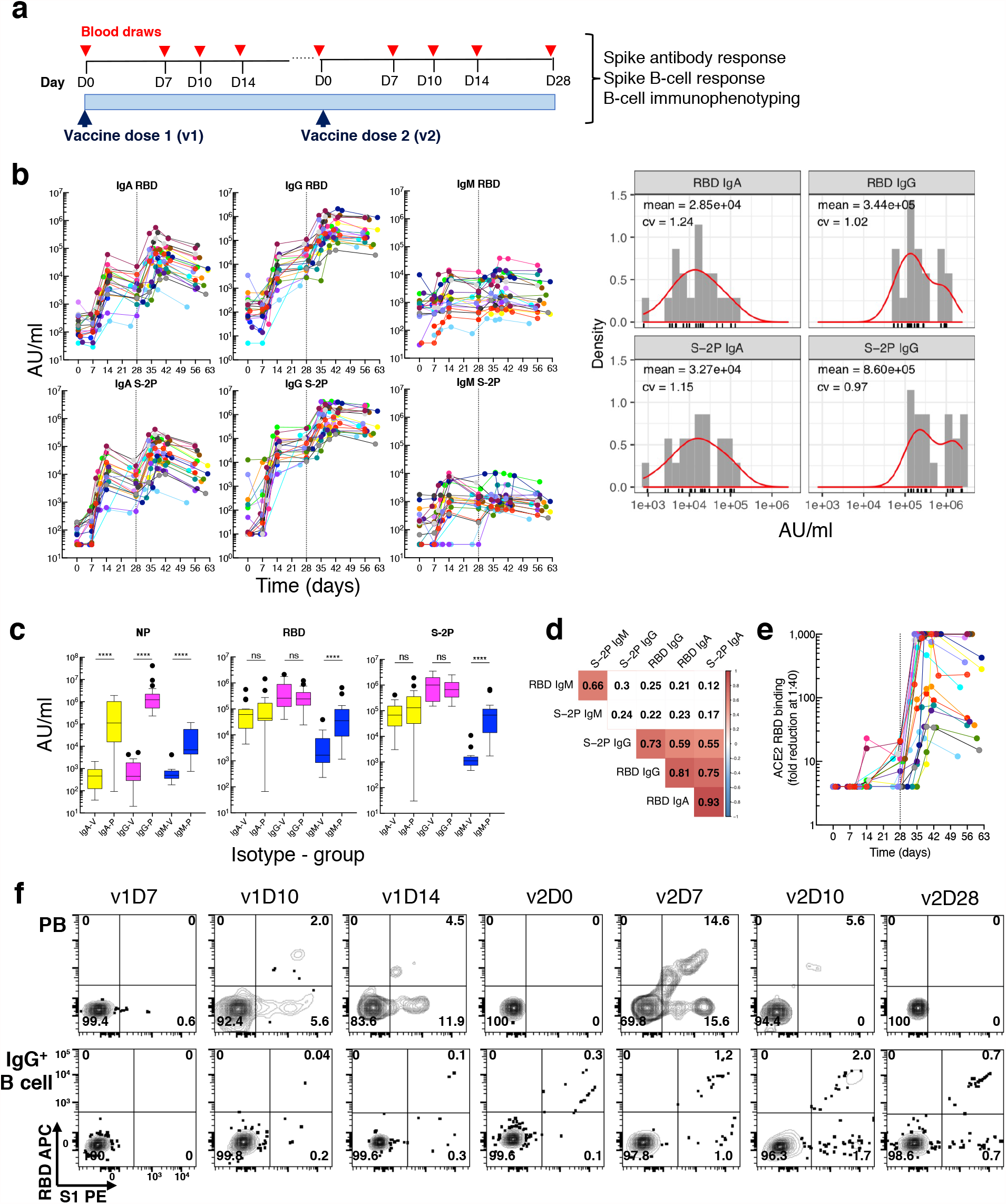
Longitudinal blood sampling and analysis shows robust antibody and early B cell response to mRNA-1273 vaccine. **a**, Study design with serial blood draws and assays performed at all timepoints on SARS-CoV-2-uninfected vaccinees (*n* = 21; missed visits in Extended Data Table 1) receiving two doses of the mRNA-1273 vaccine. **b**, Serum IgG, IgA and IgM binding to S-2P and RBD proteins measured by electrochemiluminescence (ECLIA) longitudinally (left panels), and corresponding histogram and distribution (based on kernel density estimates) at the last timepoint (v2D28) (right panels). **c**, Peak serum IgG, IgA and IgM binding to S-2P, RBD and N proteins measured by ECLIA in vaccinees (V; *n* = 21) and COVID-19 patients (P; *n* = 21), shown as boxplots. **d**, Triangular heatmap of correlation between serum antibodies at last measured timepoint (v2D28) in **(b)**. Numbers represent r values. Statistically insignificant correlations (p > 0.05) shown in white. **e**, Longitudinal inhibition of RBD binding to ACE2 by serum (1:40 dilution) of vaccinees (*n* = 21). **f**, Longitudinal binding of S1 and RBD tetramers to PB and IgG^+^ B cells by flow cytometry shown for a high responder (VAC-611; Extended Data Table 1). Numbers in each quadrant are percentages. Each vaccinee is color-coded and second vaccine dose indicated by vertical dotted line **(b**,**e)**. Mann-Whitney test; ****, p < 0.0001 **(c)**. Spearman’s rank correlation **(d)**. AU, arbitrary units; D, day; N, nucleocapsid; ns, not significant; P, patients with severe COVID-19; PB, plasmablasts; RBD, receptor binding domain; S1, spike subunit 1; S-2P, stabilized spike trimer; v, vaccine dose; V, vaccinees.

In general, B cell responses to vaccination are detected in the peripheral blood in two distinct phases; the first consisting of a short burst of PB, typically detected around D7, followed by a slower phase that leads to the establishment of a pool of long-lived MBC^18^. Antigen-specific B cells can be identified by flow cytometry using protein tetramers; this is an approach that has been used to track SARS-CoV-2 spike-specific responses following COVID-19 infection or vaccination^15,19-21^. We used a pair of RBD and spike subunit 1 (S1) tetramers to track spike-specific B cell responses of vaccinees, and as expected, dual RBD^+^ S1^+^ and single S1^+^ PB became detectable at v1D10 while corresponding MBC became detectable at v1D14 (Fig. 1f and Extended Data Fig. 1b). S-2P tetramers also clearly detected RBD^+^ within S1^+^ and S1^+^ within S-2P^+^ B cells, but they did not clearly identify S-2P^+^ PB (Extended Data Fig. 1c). We thus focused on RBD and S1 tetramers to simultaneously measure spike-specific responses among all B cell populations. We validated the approach for PB by showing that 1), frequencies of RBD^+^ and S1^+^ PB measured by flow cytometry were strongly correlated to those measured by the conventional enzyme-linked immunospot (ELISpot) assay (Extended Data Fig. 1d); and 2), IgG PB could be detected without the need for permeabilization, as is required by other methods^22^. Over 95% of PB isotypes were detected in the absence of permeabilization and their distribution was largely similar in the presence or absence of permeabilization (Extended Data Fig. 1e).

B cells that circulate in the peripheral blood are highly heterogeneous, especially MBC; their phenotypes and functions vary with the source and chronicity of antigen exposure^23^. Accordingly, we comprehensively evaluated B cells with a flow cytometric panel of 15 antibodies against primarily B cell markers and two spike tetramers (Extended Data Table 2). Unsupervised clustering analysis of the 15 cell-surface markers on CD19^+^ single B cells, performed on all vaccinees and all time-points simultaneously, revealed 30 clusters grouped by eight major B cell populations, including naïve, PB, and MBC subsets (Fig. 2a; also Extended Data Fig. 2a for more granular depictions of individual clusters, and Extended Data Table 3 for detailed annotations). A corresponding mean fluorescence intensity (MFI) heatmap delineated the surface marker phenotype, isotype, and specificity of individual cell clusters (Fig. 2b), and identified naïve B cells (C1) as the most abundant cluster, as expected for peripheral blood^22^. Five populations of conventional (CD27^+^ CD20^+^ CD21^+^) MBC, each distinguished by isotype: IgA^+^ (C14), IgG^+^ (C2 and C4) and IgD/M (C19), were prevalent. Both IgG (C9) and IgA (C13) PB were identified, as well as eight nonconventional MBC with lower abundance (Fig 2b; Extended Data Table 3 for details). IgG PB (C9) contained the highest proportion of RBD^+^ S1^+^ cells, followed by IgA PB (C13), and the nonconventional IgG^+^ MBC C5 (Fig. 2b). Our longitudinal tracking of vaccinees revealed that RBD^+^ S1^+^ PB were first detected at v1D10, then subsided until v2D5-7, while other RBD^+^ S1^+^ B cells (namely MBC) became visible at v1D14 and intensified significantly following v2 (Fig. 2c, d), consistent with the analyses performed with manual gating (Fig. 1f and Extended Data Fig. 1b).

**Fig. 2.**
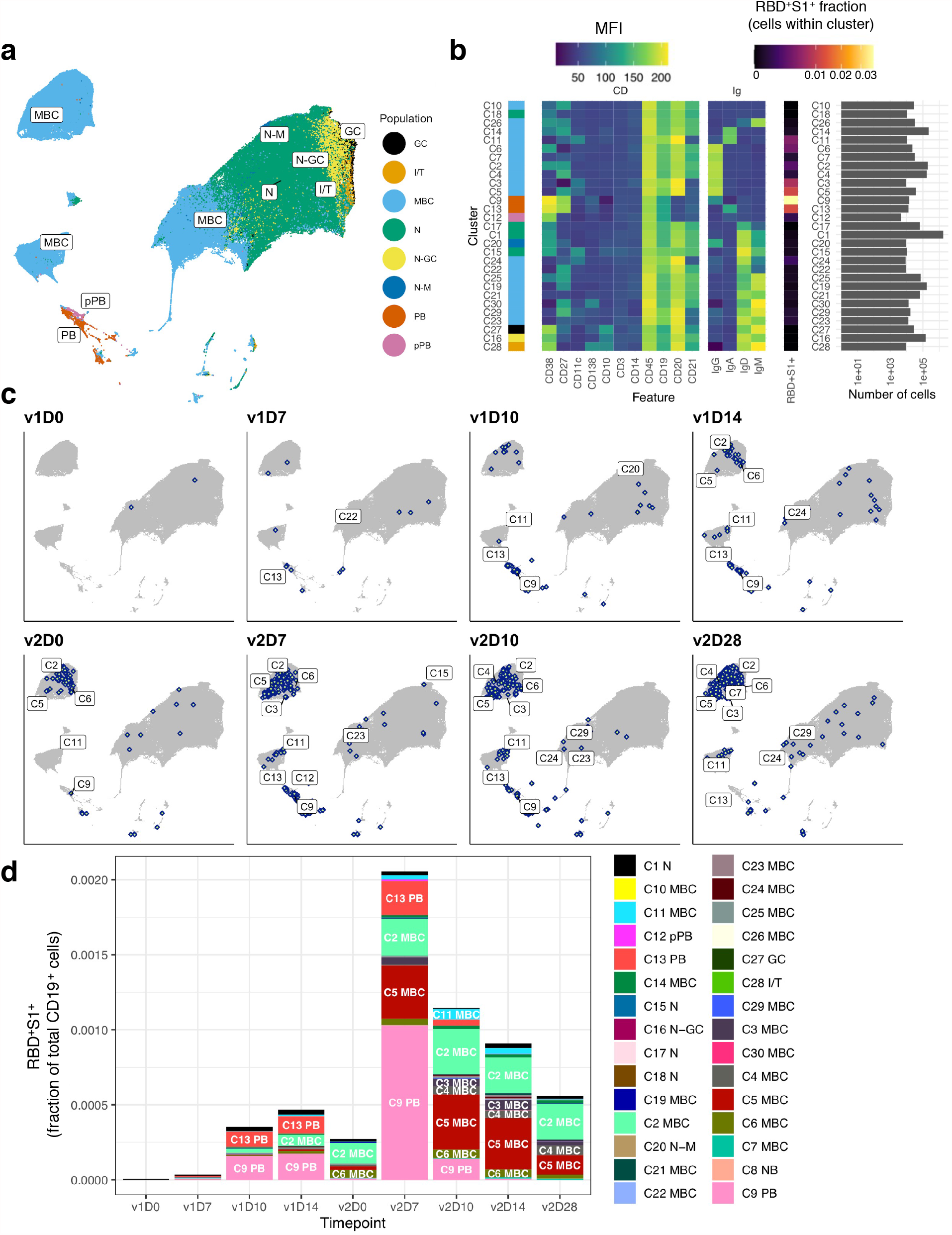
Unsupervised clustering analysis identifies major B cell populations and SARS-CoV-2-specific B cells. **a**, UMAP projection of combined B cells (n = 653,683 cells), subsampled from 3.2 million CD19^+^ cells to include 3,667 cells per sample and all RBD^+^ S1^+^ cells from all study participants (*n* = 21) at all timepoints with annotated major B cell populations identified by FlowSOM clustering. **b**, MFI-based heatmap of FlowSOM clusters as indicated by cluster number and marker. Rows ordered by hierarchical clustering. Summary of fraction of cells binding both RBD and S1 within each cluster and cell counts per cluster (right). **c**, UMAP plots with overlays of RBD^+^ S1^+^ B cells (blue points with white center) at each timepoint. **d**, RBD^+^ S1^+^ cells within each cluster expressed as a fraction of total CD19^+^ B cells across all subjects at each timepoint (*n* at each timepoint shown in Extended Data Table 1). D, day; GC, germinal center; I/T, immature transitional; MBC, memory B cell; MFI, mean fluorescence intensity; N, naïve; PB, plasmablast; pPB, pre-plasmablast; v, vaccine dose.

We next used a linear model to search for cell clusters (both antigen non-specific and specific) whose frequencies varied significantly as a function of time in response to vaccination (Fig. 3a). Antigen non-specific clusters exhibited distinct patterns of response kinetics (Fig. 3b, c). Most notable were the decreasing nonconventional CD27^-^ IgG^+^ MBC C3 and C6 after v1 and v2, while the two PB, IgG C9 and IgA C13, and the IgA pre-PB C12 were sharply increasing after each dose. Other temporally changing clusters included several MBC and GC founder B cells (C27) that underwent modest changes after v1 and v2 (Fig. 3b, c), possibly a reflection of trafficking to and from lymphoid tissues.

**Fig. 3.**
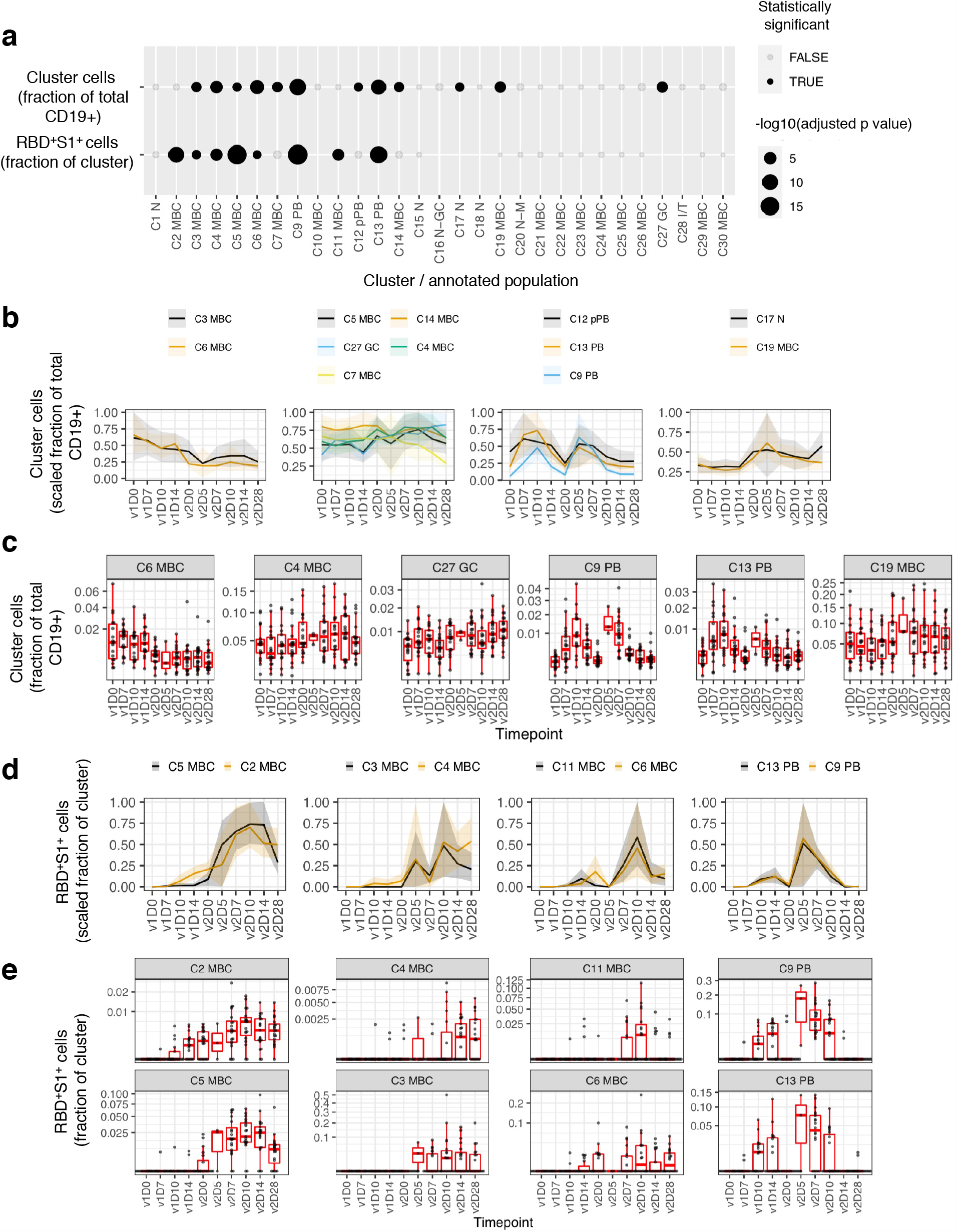
Antigen non-specific and spike-specific cells exhibit temporal change in response to the mRNA-1273 vaccine. **a**, Clusters showing significant temporal variation over course of v1 and v2 in the frequency of non-specific cells as a fraction of total (CD19^+^) B cells (first row) and RBD^+^ S1^+^ cells as a fraction within each cluster (second row). **b**, Longitudinal display of non-specific cells per cluster as a fraction of total (CD19^+^) B cells, shown for clusters with statistically significant temporal variations, as shown in **(a)**. Clusters were grouped by temporal patterns (see methods). Lines denote the mean and shading denotes 95% bootstrap confidence interval per timepoint. Values rescaled as fraction of maximum 95% confidence interval estimate over the entire time course. **c**, Similar to **(b)** but showing boxplots for selected clusters. **d**, similar to **(b)** but displaying of RBD^+^ S1^+^ cells as a fraction within each cluster. **e**, Similar to **(d)** but showing boxplots. Type III ANOVA test using Satterthwaite’s approximation **(a)**. N at each timepoint shown in Extended Data Table 1 **(b-e)**. D, day; RBD, receptor binding domain; S1, spike subunit 1; v, vaccine dose.

Spike-specific B cell frequencies, measured as a fraction of RBD^+^ S1^+^ cells among cells within each cluster, also exhibited varying patterns of temporal responses (Fig. 3d). Two-peak responses with stronger increases in v2 than v1 were observed for the two PB clusters C9 and C13, nonconventional MBC C11 (CD27^lo^ IgA^+^) and C6 (CD27^-^ IgG^+^), albeit with differences in timing. For example, C9 (IgG) and C13 (IgA) PB had an initial modest burst beginning at v1D10, followed by a second stronger but shorter burst at v2D5-7 (Fig. 3e). RBD^+^ S1^+^ cells among two pairs of conventional/nonconventional MBC, namely C2/C5 and C3/C4 respectively, also underwent coordinated changes following v2 (Fig. 3d). It is notable, however, that while RBD^+^ S1^+^ cells in all four of these MBC clusters showed trends of declines from their peaks by v2D28, those in the two nonconventional MBC (C3 and C5) appeared to drop more precipitously than in the two conventional MBC (C2 and C4; Fig. 3e). These findings are consistent with recent reports that spike-specific memory responses several months after SARS-CoV-2 infection are enriched within CD21^+^ CD27^+^ MBC^17,21^, which have a phenotype similar to C2 and C4.

Despite the coherent changes observed across subjects, substantial heterogeneity in B cell responses existed among vaccinees. Independent of antigen specificity, the magnitude of PB increases is known to be a correlate of antibody responses for vaccines such as influenza^24,25^. We thus assessed associations between the changes in non-specific cluster frequencies over the course of v1 and v2 relative to the respective baselines with IgA and IgG RBD and/or S-2P titers at v2D28 by using linear models accounting for age and gender (Extended Data Fig. 3a, b). Both IgA and IgG PB (C9 and C13) at v1D10 were indeed positively associated, albeit mildly, with both RBD and S-2P IgA titers (but not IgG; Extended Data Fig. 3a, c, d), while several MBC clusters (C3, C11, C24) at as early as v1D7 and v1D10 were correlated with S-2P IgG titers (Extended Data Fig. 3a).

The frequency of spike-specific PB on v2D7 spanned a wide range (Fig. 3e), raising the question of whether those with depressed PB responses also had lower antibody titers following v2. Thus, we next used the same linear model to search for spike-specific correlates (Fig. 4a, b). Indeed, at v2D7 and v2D10, IgG PB (C9) were correlated with IgG and IgA RBD antibodies, while IgA PB (C13) at v2D7 were associated with IgA S-2P antibodies (Fig. 4b, c). Thus, in contrast to antigen non-specific PB, spike-specific PB appeared as serological correlates only after v2. However, the frequency of RBD^+^ S1^+^ cells within IgA PB C13 at v1D10 predicted the fraction of RBD^+^ S1^+^ cells in the C2 MBC at v2D7 (Extended Data Fig. 3e), suggesting that the magnitude of the spike-specific PB expansion after v1 is associated with the memory response after v2. A recent preprint did not find a correlation between antibody response and transcriptional modules enriched for PB at D7 following the second dose of the Pfizer vaccine^26^, likely due to differences in assessing PB responses using blood transcriptional signatures versus our direct measurement of fresh, antigen-specific PB. In addition to PB, spike-specific C6 was a positive correlate at v2D0 (Fig. 4a, d) when these cells reached a first peak (Fig. 3d). C6 are CD27^-^ IgG^+^ MBC known to have lower mutational burdens than their CD27-expressing counterparts^27^, and may as such, reflect products of early events of the antigen-driven maturation process after v1. C6 are also CD21^hi^, and similar to a stable pool of CD27^-^ MBC that are generated in response to new influenza variants^28^.

**Fig. 4.**
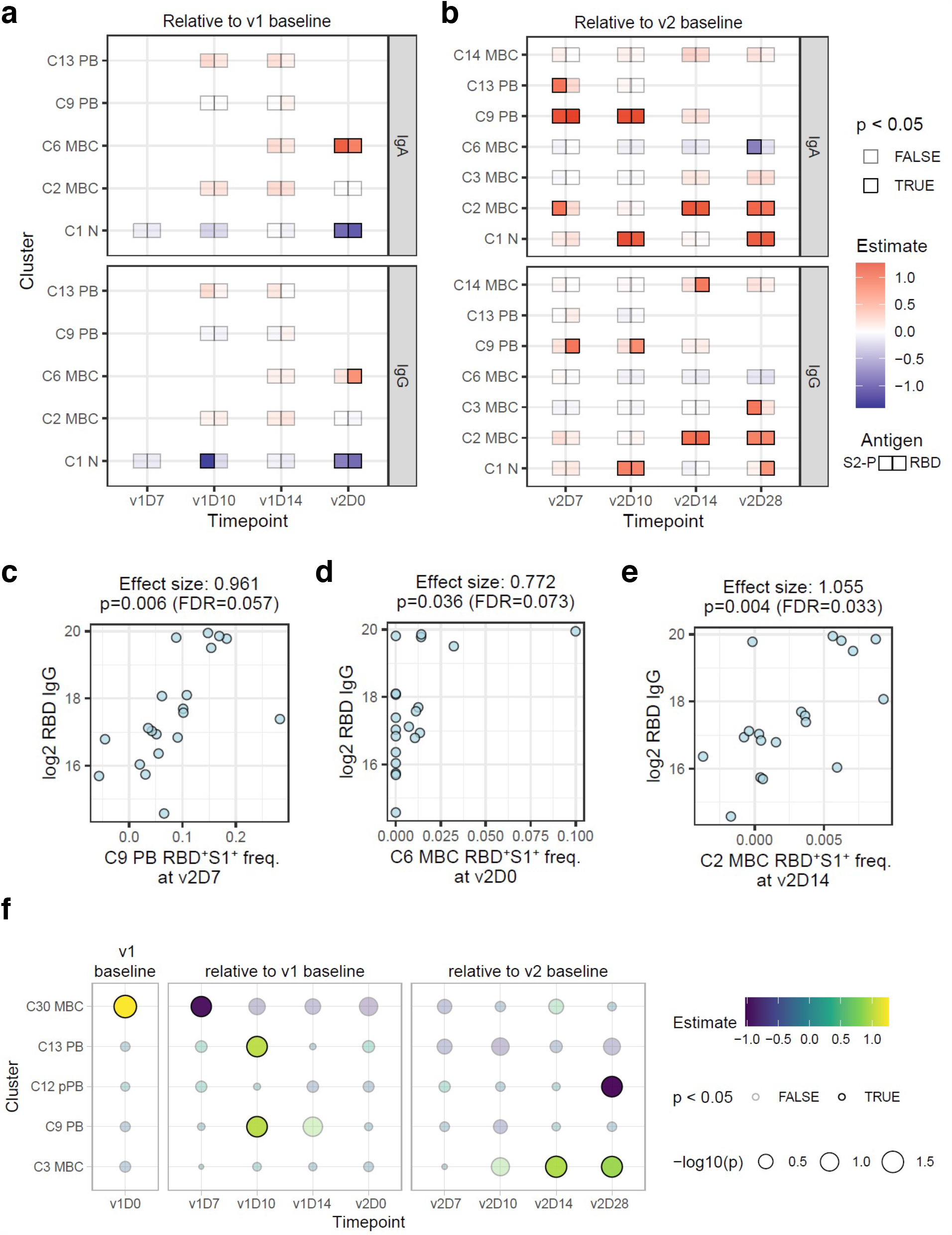
Correlates of SARS-CoV-2 antibody titers 28 days after second dose of vaccine. **a,b**, Linear model effect size estimates indicate strength of association between spike-specific (RBD^+^ S1^+^) cell frequency in the cell clusters (rows) with antibody endpoints (IgA and IgG titers for S-2P and RBD) relative to pre-vaccination baseline level (v1D0) at four timepoints between v1 and v2 **(a)**, and relative to v2 baseline (v2D0) at four timepoints after v2 **(b)**. Only clusters with at least one significant (unadjusted p ≤ 0.05) association at any timepoint are shown. At each timepoint, clusters that had fewer than five samples with any RBD^+^ S1^+^ cells were excluded from analysis (missing boxes). **c-e**, Scatter plots illustrating correlations between endpoint (v2D28) RBD IgG titers and RBD^+^ S1^+^ cell frequencies in C9 on v2D7, C6 on v2D0, and C2 on v2D14, respectively. Effect sizes and p values were estimated by the linear models above. FDR estimate of the statistical significance was calculated within each antibody endpoint and timepoint combination. **f**, Effect size estimates of association between first principal component (PC1) of endpoint SARS-CoV-2 antibody titers and frequencies of each cell cluster as a fraction of total CD19^+^ cells. PC1 was derived from IgA and IgG titers against S-2P and RBD proteins at v2D28. Only cell clusters with at least one significant (unadjusted p ≤ 0.05) association at any timepoint are shown. D, day; FDR, false discovery rate; RBD, receptor binding domain; rho, Spearman’s rank correlation; S1, spike subunit 1; S-2P, stabilized spike trimer; v, vaccine dose.

Most of the other antigen-specific correlates were positively associated with the titer response and reflect changes after the second dose (Fig. 4b), including spike-specific MBC. At v2D14 and v2D28, the frequency of RBD^+^ S1^+^ cells in conventional MBC C2 was positively correlated with RBD/S-2P IgA and IgG antibodies (Fig. 4b, e), consistent with their role in sustaining immunological memory^17,21^. In contrast, it is notable that C5, the nonconventional MBC with features of vaccine-induced activated MBC^24^, and that we found to contain a strong RBD/S1 response following v2 (Fig. 3d, e), did not correlate with endpoint antibodies (data not shown). Together, these may reflect a unique response feature of this mRNA vaccine, but timing or insufficient statistical power may have played a role in the lack of detection of correlation between C5 and antibodies.

Among the clusters that correlated with antibodies, several were not of the same isotype. While it is possible that IgG B cells could give rise to IgA-secreting cells^29^, the reverse cannot occur. Thus, the most likely explanation for inter-isotype correlations is that most are not causal, but reflect a coordinated immunologic response driven by shared mechanisms, as indicated by the strong correlations between IgG and IgA antibodies (Fig. 1d). Indeed, when we used the correlated component (the first principal component) of the v2D28 IgG and IgA RBD and S-2P antibody titers as an isotype-independent endpoint, we found many of the positive antigen-specific correlates highlighted above, including C6 MBC at v2D0, C9 PB at v2D7, and C2 MBC at v2D14 (Extended Data Fig. 3f), indicating that these correlates reflected isotype independent responses that potentially determined the magnitude of antibodies induced by the mRNA vaccine.

Intriguingly, aside from confirming the early plasmablast correlates at v1D10 above, the isotype independent analysis revealed additional non-specific correlates from as early as the baseline before the first dose of vaccination (Fig. 4f): C30 MBC was a positive baseline predictor independent of age and gender, suggesting that the frequency of these circulating unswitched CD138^+^ MBC reflected an antigen non-specific “set point” for humoral responses to naïve antigens^8^. Consistent with this notion, C30 did not appear again as a baseline (v2D0) correlate for the second dose (data not shown), probably because v2 elicited a recall response. Interestingly, by v1D7, this population was a negative correlate (Fig. 4f), perhaps because its lower cell frequency in blood by then reflected greater ongoing B cell activation and differentiation into class-switched PB and MBC. Together, our analyses revealed both antigen non-specific and spike-specific predictive signatures of v2D28 antibody responses. These signatures could help predict and monitor the serological efficacy of SARS-CoV-2 mRNA vaccines and pave the way to a better understanding of weakened responses to this novel vaccine platform, such as those associated with age and chronically compromised immune systems.

## Methods

### Study design and participants

Twenty SARS-CoV-2-uninfected NIH employees and one community member who were eligible to receive an Emergency Use Authorization COVID-19 vaccine were recruited to study longitudinal vaccine responses. The 21 participants received a first dose of the Moderna mRNA-1273 vaccine and 25 to 34 days later, 20 participants received a second dose of the same vaccine (Extended Data Table 1). One participant was lost to follow-up after testing positive for COVID-19 between doses. To evaluate differences in SARS-CoV-2 antibody responses between vaccination and infection, we recruited twenty-one patients with severe COVID-19 (Extended Data Table 4). An additional 11 participants who received both doses of the Moderna mRNA-1273 vaccine were recruited for the validation analyses of plasmablast (PB) frequencies between flow cytometry and ELISpot; nine were SARS-CoV-2-uninfected and two had recovered from COVID-19 infection. Research phlebotomy was performed at the NIH Clinical Research Center in Bethesda, MD under protocols approved by the NIH Institutional Review Board, ClinicalTrials.gov identifiers: NCT00001281, NCT04411147, NCT04280705, and NCT04579393. All participants provided written informed consent.

### Blood sample collection and processing

Peripheral blood mononuclear cells (PBMC) were isolated by Ficoll density gradient centrifugation from whole blood collected in EDTA vacutainer tubes. Serum was isolated by centrifugation of clotted whole blood collected in SST vacutainer tubes and stored at -80°C.

### SARS-CoV-2-binding antibody assay

Serum samples were heat inactivated at 56°C for 60 minutes. A 4-plex antibody binding assay was performed using an electrochemiluminescence immunoassay analyzer (ECLIA) developed by Meso Scale Discovery (MSD). Each well of MSD SECTOR^®^ plates was precoated by manufacturer (MSD) with SARS-CoV-2 spike (S-2P), receptor binding domain (RBD) protein, nucleocapsid (N) protein and a Bovine Serum Albumin (BSA) in a specific spot-designation for each antigen. Plates were blocked at room temperature (RT) for 60 minutes with MSD blocker A solution containing 5% BSA. Plates were washed and MSD reference standard (calibrator), QC test sample (pool of COVID-19 convalescent sera) and human serum test samples were added in duplicate in an 8-point dilution series and reference standards were added in triplicate. MSD Control sera (low, medium and high) were added undiluted in triplicate. Samples were incubated with shaking at RT for 4 hours on a Titramax Plate shaker (Heidolph). Plates were washed and incubated with MSD SULFO-TAG™ anti-human IgG, IgA or IgM detection antibodies at RT for 60 minutes with shaking. Plates were washed, MSD GOLD™ read buffer containing electrochemiluminescence (ECL) substrate was added, and plates were read with the MSD MESO Sector S 600 detection system. Analyses were performed with Excel (Microsoft) and Prism 9.0 (Graphpad) software and antibody concentrations were assigned arbitrary units (AU/ml) by interpolation from the standard curve. Densities of antibody concentrations at endpoint (v2D28) were estimated using a gaussian kernel with bandwidth automatically selected through biased cross validation using the stat_density function from ggplot2 (3.3.3) with bw = “bcv”.

### RBD-ACE2 blocking assay

Samples were prepared as for the 4-plex binding assay. 384-well plates precoated with RBD were supplied by the manufacturer (MSD). Plates were blocked at room temperature (RT) for 30 minutes with MSD blocker A solution containing 5% BSA. Plates were washed, test samples were added at dilutions of 1:10, 1:20 and 1:40, and incubated with shaking at RT for 60 minutes. Human ACE2 conjugated with SULFO-TAG was added and plates were further incubated to allow binding to RBD. Plates were washed, ECL substrate added, and plates read as in antibody binding assay. Fold reduction in ECL response for each sample was calculated against based on signal emitted in wells in absence of sample (assay diluent).

### Recombinant biotinylated RBD protein

A SARS-CoV-2 RBD construct containing a His-tag and Avi-tag was generated, as previously described^30^. The residues 319-541 of the S protein were codon optimized with N-terminal of signal peptide (MFVFLVLLPLVSSQ) and C-terminal of 6-His tag and Avi-tag (GLNDIFEAQKIEWHE). The DNA encoding sequence was cloned into the mammalian cell expression vector pCAGGS and confirmed by sequencing, prior to transient transfection in FreeStyle 293-F cells with 293fectin transfection reagent (ThermoFisher). Culture supernatants were harvested at 5 days post transfection, filtered, and purified by in-house packed affinity purification column with Complete His-tag purification resin (Roche). Elutes were buffer exchanged with phosphate-buffered saline (PBS), and concentrated using an Amicon Ultra 10 kDa molecular weight cutoff concentrator (Millipore). Biotinylation was performed with a BirA biotin-protein ligase standard reaction kit (Avidity), according to the manufacturer’s instructions. Excess biotin was removed by five buffer exchanges with an ultra 10K concentrator (Amicon).

### B-cell spike-specific responses and phenotyping by spectral flow cytometry

A 17-color panel was developed to phenotype B-cell populations and identify SARS-CoV-2-specific B cells among PBMC (Extended Data Table 2 for list and source of antibodies and biotinylated spike proteins). The biotinylated spike proteins were tetramerized with fluorescently labeled streptavidin (SA) as follows: S1 with SA-R-Phycoerythrin (PE), RBD with SA-Allophycocyanin (APC), and S-2P with SA-Alexa Fluor 488 (Thermo Fisher Scientific). In a stepwise process, 1/5 of the molar equivalent of the SA-fluorochrome reagent was added to the biotinylated protein at 20-min intervals until the molar ratio of biotinylated protein and streptavidin-fluorochrome reached 4:1. Incubations were carried out at 4°C with gentle rocking. To titrate the labeled protein tetramers and establish background and antigen-specificity, freshly isolated or cryopreserved PBMC from SARS-CoV-2-uninfected and recovered infected individuals were used as negative and positive controls, respectively. For vaccinees, 10^6^ freshly isolated PBMC were stained with a cocktail containing the 15 panel antibodies and 160 ng each of PE-conjugated S1 and APC-conjugated RBD in staining buffer (2% FBS/PBS) supplemented with Brilliant Stain Buffer Plus (BD Biosciences) at 4°C for 30 minutes. In a second 18-color panel, 400 ng of Alexa Fluor 488 conjugated S-2P was added to the cocktail. The stained cells were acquired on an Aurora spectral cytometer (Cytek Biosciences) and analyzed using FlowJo v10.7.1 (BD Biosciences).

### Intracellular flow cytometry

PBMC were first stained with antibodies against cell surface markers CD3, CD19, CD20 and CD27, fixed (Lysing Solution, BD Biosciences), permeabilized (Permeabilizing Solution 2; BD Biosciences) and stained with antibodies against IgG, IgA, IgD and IgM (antibody details in Extended Data Table 5). The stained cells were acquired on a FACS Canto II flow cytometer (BD Biosciences) and analyzed using FlowJo software v9.9.6 (BD Biosciences).

### ELISpot assay to enumerate SARS-CoV-2 spike-specific PB

Spike protein S1 and RBD-specific antibody-secreting PB were enumerated by modifying the antigen-specific portion of a previously described ELISpot assay^31,32^. Briefly, wells of Immobilon-P polyvinylidene difluoride (PVDF) membrane 96-well plates (Millipore) were coated with 5ug/ml anti-Ig light-chain antibodies (Rockland Immunochemicals) overnight at 4°C. Plates were washed and wells were blocked at RT for 2 hours with RPMI containing 10% FBS. Duplicate wells were plated with PBMC containing 0.01-0.003 × 10^6^ B cells for total IgA/G/M-secreting PB enumeration and 0.1-0.03 × 10^6^ B cells for RBD/S1-specific PB. Plates were incubated at 37°C for 5 hours, followed by overnight incubation at 4°C with biotinylated antibodies (Jackson Immunoresearch) against IgA (catalogue 109-066-011), IgG (catalogue 709-066-149), IgM (catalogue 709-066-073), or biotinylated proteins S1 (Acrobiosystems, catalogue S1N-C82E8) or RBD (Biolegend, catalogue 790904). Plates were washed, streptavidin-AP conjugate (R&D Systems) was added, followed by incubation at RT for 2 hours. Plates were washed and spots were developed with ELISpot Blue Color Module (R&D Systems).

Biotinylated (Biorbyt) or unlabeled (Millipore-Sigma) keyhole limpet hemocyanin (KLH) was used as negative control antigen to enumerate background spots. Spots were counted using an ELISPOT reader (Cellular Technology Ltd). Frequencies of S1 and RBD-specific PB were calculated as the fraction of total Ig-secreting PB after subtraction of background KLH spots.

### Spectral flow cytometry data processing for FlowSOM clustering and UMAP embedding

FCS files generated from spectral flow cytometry with the 17-color panel and associated manual gates were read into R using FlowWorkspace (4.2.0). Live, CD19^+^ single cells were selected for downstream analysis. Data from all samples were merged and transformed with the arcsinh transformation with a scale factor of 1/150. Data were not z-score scaled. The following markers were used to perform clustering CD20, CD138, CD38, CD10, CD11c, CD19, CD27, CD21, IgD, IgM, IgG, IgA, using FlowSOM (1.22.0)^33^, with the number of desired metaclusters (nClus) set to 30. One small cluster, C8, determined to represent granulocytes, was removed from downstream analysis. To visualize the clusters and RBD^+^ S1^+^ cells in a UMAP embedding^34^, 653,683 cells were subsampled from the roughly 3.2 million CD19^+^ cells to include 3,667 cells per sample and all RBD^+^ S1^+^ cells from the 21 longitudinal vaccine participants at all timepoints. Data were transformed as described above and the UMAP embedding was fit using the uwot package (0.1.10) with default parameters.

### Analysis of temporally varying FlowSOM clusters

Frequencies of cells within each sample were summarized into 1) the fraction of cells within a cluster relative to the total number of CD19^+^ cells in the given sample; and, 2) the fraction of cells within a cluster determined to be RBD^+^ S1^+^ by manual gating, relative to the number of cells in that cluster in the given sample. The extent of temporal variation was assessed using a linear mixed effects model with the following formula in lme4 (1.1.26):

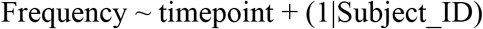

Timepoint is a factor variable representing the discrete timepoints (v1D0, v1D7, etc.)

The significance of the timepoint term was assessed with a type III ANOVA using Satterthwaite’s approximation using the lmerTest package (3.1.3)^35^. P values were adjusted across all comparisons using the Benjamini-Hochberg procedure^36^. Clusters with adjusted p values below 0.05 were deemed temporally fluctuating. Clusters selected by the above procedure were then grouped by the similarity of their temporal patterns. Briefly, the mean frequency across all subjects at a given timepoint was computed along with the 95% bootstrap confidence intervals around the mean using package Hmisc (4.5.0). The means were then rescaled by dividing all values by the maximum value of the 95% confidence interval throughout the time-course such that all values were now in the range of [0,1]. The clusters were then grouped by hierarchical clustering of the mean trends using the Euclidean distance at each timepoint and using Ward’s method, as implemented in the hclust function (method = “ward.D2) in R (4.0.2). After inspecting the respective dendrograms, 4 groups were determined to be appropriate, and the hierarchical clustering trees were cut to produce 4 groups for both antigen non-specific and specific cells.

### Modeling of association between endpoint antibody concentrations and cluster frequencies

A linear model accounting for the age and gender of the longitudinal vaccine participants was used to estimate whether cell cluster frequencies in response to vaccination were associated with SARS-COV2 spike protein (S-2P/RBD) antibody concentration at endpoint (v2D28):

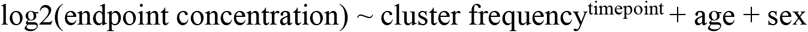

Analyses were carried out on both standardized antigen non-specific and specific frequencies, i.e., cluster cell counts as a fraction of total CD19^+^ cell counts and RBD^+^ S1^+^ cells within the clusters, respectively. For the antigen non-specific models, only clusters whose post-vaccination frequencies at any timepoint changed significantly from the pre-vaccination baseline (v1D0) were included. For the antigen-specific models, clusters with at least four RBD^+^ S1^+^ cells in any of the samples were considered. In addition, at each timepoint, a cluster was excluded if there were fewer than five samples with any RBD^+^ S1^+^ cells. Pre-vaccination baseline frequencies (v1D0) were subtracted from frequencies post vaccine dose 1 (v1) timepoints, including v1D7, v1D10, v1D14, and v2D0. Similarly, dose 2 baseline frequencies (v2D0) were subtracted from frequencies of post-vaccine dose 2 timepoints, including v2D7, v1D10, v1D14, and v2D28. P values were adjusted with the Benjamini-Hochberg method within each combination of timepoint and antibody endpoint^36^. R version 3.6.3 was used for this analysis.

### Correlation between principal component of endpoint antibody concentrations and cluster frequencies

In addition to modeling the association of the cluster frequencies to individual antibody concentrations, their relationship to the primary correlated component of the two antibodies to both S-2P and RBD proteins was also assessed. PC1 from principal component analysis of the four endpoints (in log2 scale), i.e., S-2P IgA/G and RBD IgA/IgG explained 83.6% of variance across subjects. Associations between PC1 and RBD^+^ S1^+^ cluster frequencies at each timepoint were calculated using the same linear models and inclusion criteria described above. The same analysis was carried out for all antigen non-specific clusters (cell counts as a fraction of total CD19^+^ B cells) with the baseline pre-vaccination timepoint (v1D0) included.

## Supporting information

All supplemental materials

## Data Availability

All data are available in the main text or supplementary materials.

## Acknowledgements

The authors thank all the participants for their willingness to take part in our study; Catherine Rehm and Ulisses Santamaria for coordinating the collection and distribution of clinical samples for this study; NIAID OCICB for high-performance computing (HPC) support. This work was funded by the Intramural Research Program of the NIAID of the NIH and NIH extramural grant R01AI102766 (YL). YL was also supported by the University of Maryland Strategic Partnership (MPower).

## Author contributions

J.S.T., A.S.F., L.K, N.R., W.W.L., A.M. and S.M. designed research and methodology; L.K., N.R., W.W.L., C.M.B., S.A., S.O., S.N., K.T., F.L.D.A., W.W., X.Z. and T.W.C. performed experiments; Y.W., C.I.C., Y.L. and A.M. provided reagents and resources; L.K., N.R., W.W.L., S.O., C.M.B., T.W.C., J.S.T. and S.M. analyzed the data; N.R. and W.W.L. wrote the software; R.R., G.E.M., C.A.S., R.W.C., A.F.S., J.R.S., D.S.C., R.T.D. and M.C.S. contributed to recruitment of study participants; L.K., N.R., W.W.L., C.M.B., T.W.C., A.S.F., J.S.T. and S.M. wrote the manuscript.

## Data availability

All data are available in the main text, extended data and excel tables.

## Code availability

All codes will be made available upon publication without restrictions.

