## Supplementary material for "Pre-vaccination and early B cell signatures predict antibody response to SARS-CoV-2 mRNA vaccine": All supplemental materials

**Kardava et al. Supplementary Materials**

Extended Data Figures 103

Extended Tables 1-5

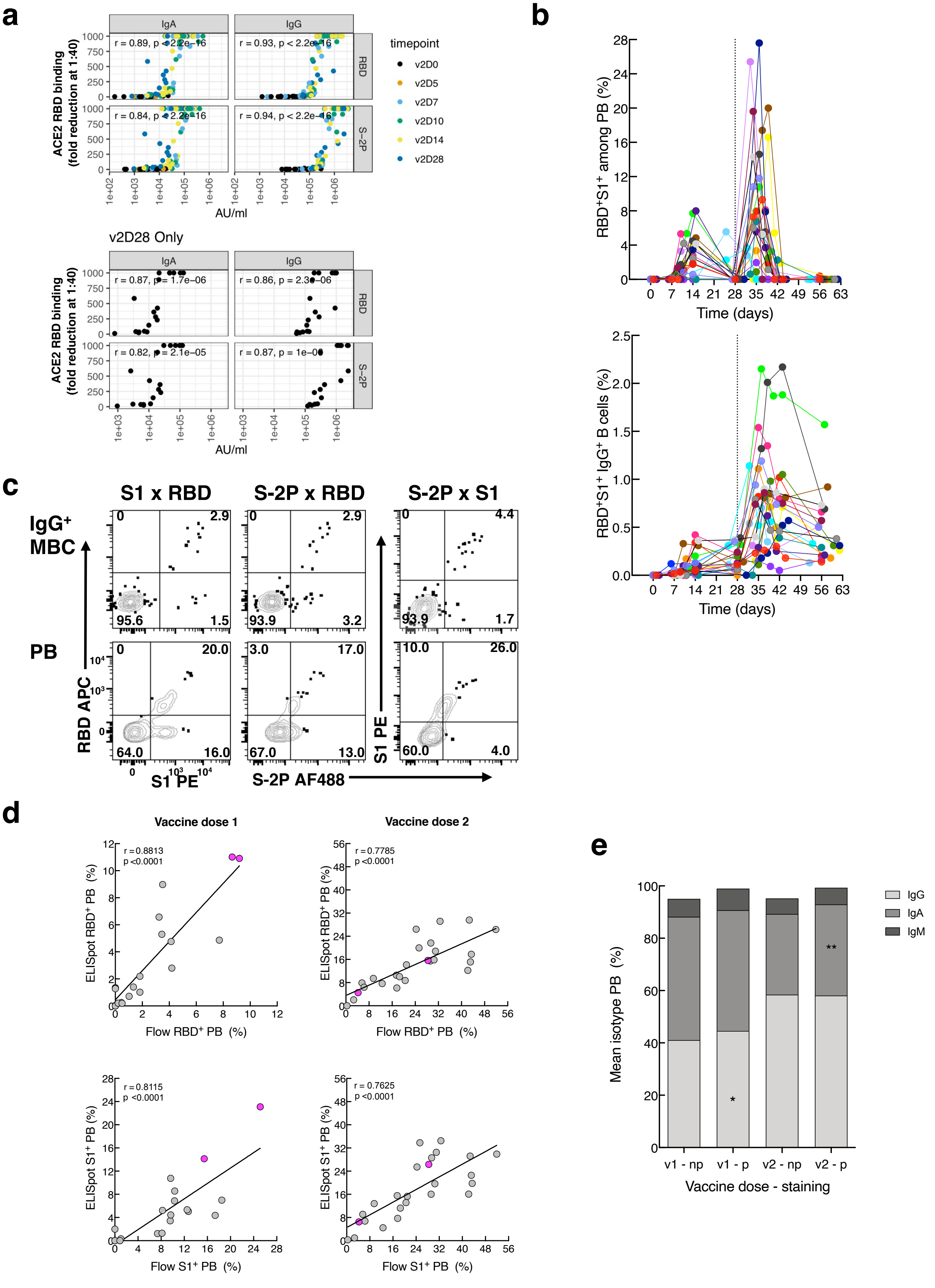
a
IgA
IgG
RBD
S−2P
r = 0.89, p < 2.2e−16
r = 0.93, p < 2.2e−16
r = 0.84, p < 2.2e−16
r = 0.94, p < 2.2e−16
ACE2 RBD binding (fold reduction at 1:40)
AU/ml
timepoint
v2D0
v2D5
v2D7
v2D10
v2D14
v2D28
v2D28 Only
r = 0.87, p = 1.7e−06
r = 0.86, p = 2.3e−06
r = 0.82, p = 2.1e−05
r = 0.87, p = 1e−06
b
RBD+S1+ among PB (%)
Time (days)
RBD+S1+ IgG+ B cells (%)
c
S1 x RBD
S-2P x RBD
S-2P x S1
IgG+ MBC
PB
RBD APC
S1 PE
S-2P AF488
d
Vaccine dose 1
Vaccine dose 2
r = 0.8813
p < 0.0001
r = 0.7785
p < 0.0001
r = 0.8115
p < 0.0001
r = 0.7625
p < 0.0001
ELISpot RBD+ PB (%)
Flow RBD+ PB (%)
ELISpot S1+ PB (%)
Flow S1+ PB (%)
e
Mean isotype PB (%)
IgG
IgA
IgM
v1 - np
v1 - p
v2 - np
v2 - p
Vaccine dose - staining

**Extended Data Fig. 1. Antibody and cellular assays with validations and correlations. a,** Correlations between serum IgG and IgA binding to RBD (Fig. 1b) and ACE2 inhibition assay (Fig. 1e) with v2 timepoints shown with color-coding (top panel) or exclusively measurements at v2D28 (lower panel). **b,** Longitudinal frequencies of $RBD^+S1^+$ PB and $IgG^+$ B cells vaccinees (*n* = 21) color-coded as in Fig. 1. **c,** Binding by flow cytometry of S-2P, S1 and RBD tetramers to PB and $IgG^+$ B cells of an individual at v2D6. **d,** Correlation between flow cytometric and ELISpot assays measuring frequencies of $RBD^+$ and $S1^+$ PB at peak dose 1 and 2 post mRNA-1273 vaccination. **e,** Average PB isotype distribution measured by flow cytometry in presence or absence of permeabilization for donors/dose timepoints in **(d)**. Donors **(d,e)** were participants in protocol NCT00001281 (n = 20) and NCT04411147 (n = 6). Of the 26 donors, two had prior SARS-CoV-2 infection (magenta circles in **(d)**). Spearman's rank correlation **(a,d)**. Paired t test.*, $p < 0.05$; **, $p < 0.01$ **(e)**. AU, arbitrary units; D, day; NP, not permeabilized; P, permeabilized; PB, plasmablasts; RBD, receptor binding domain; S1, spike subunit 1; S-2P, stabilized spike trimer; v, vaccine dose; V, vaccinees.

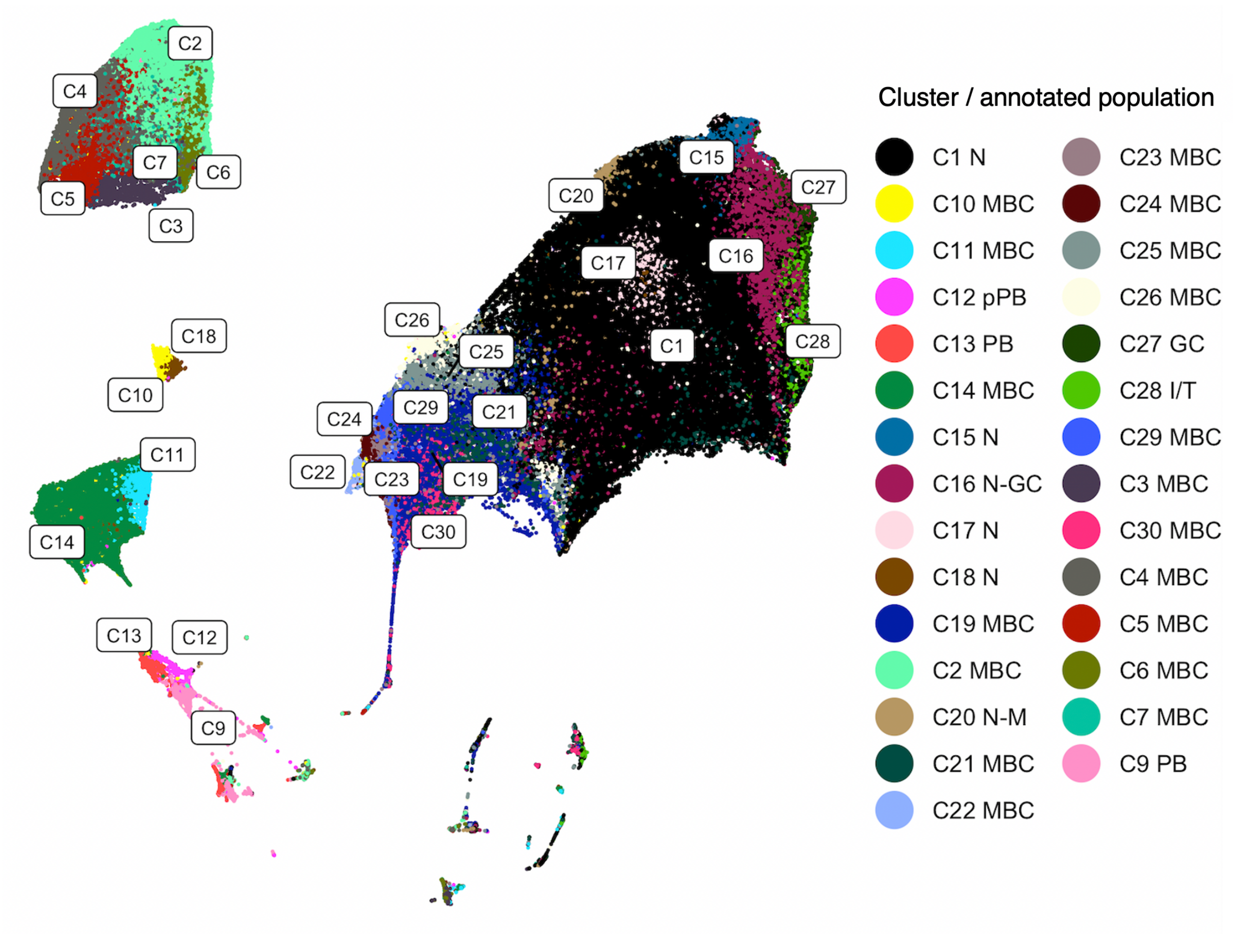


**Extended Data Fig. 2. Uniform Manifold Approximation and Projection (UMAP) displaying location of individual cell population clusters.** Similar to Fig. 2a but displaying annotations of individual clusters.

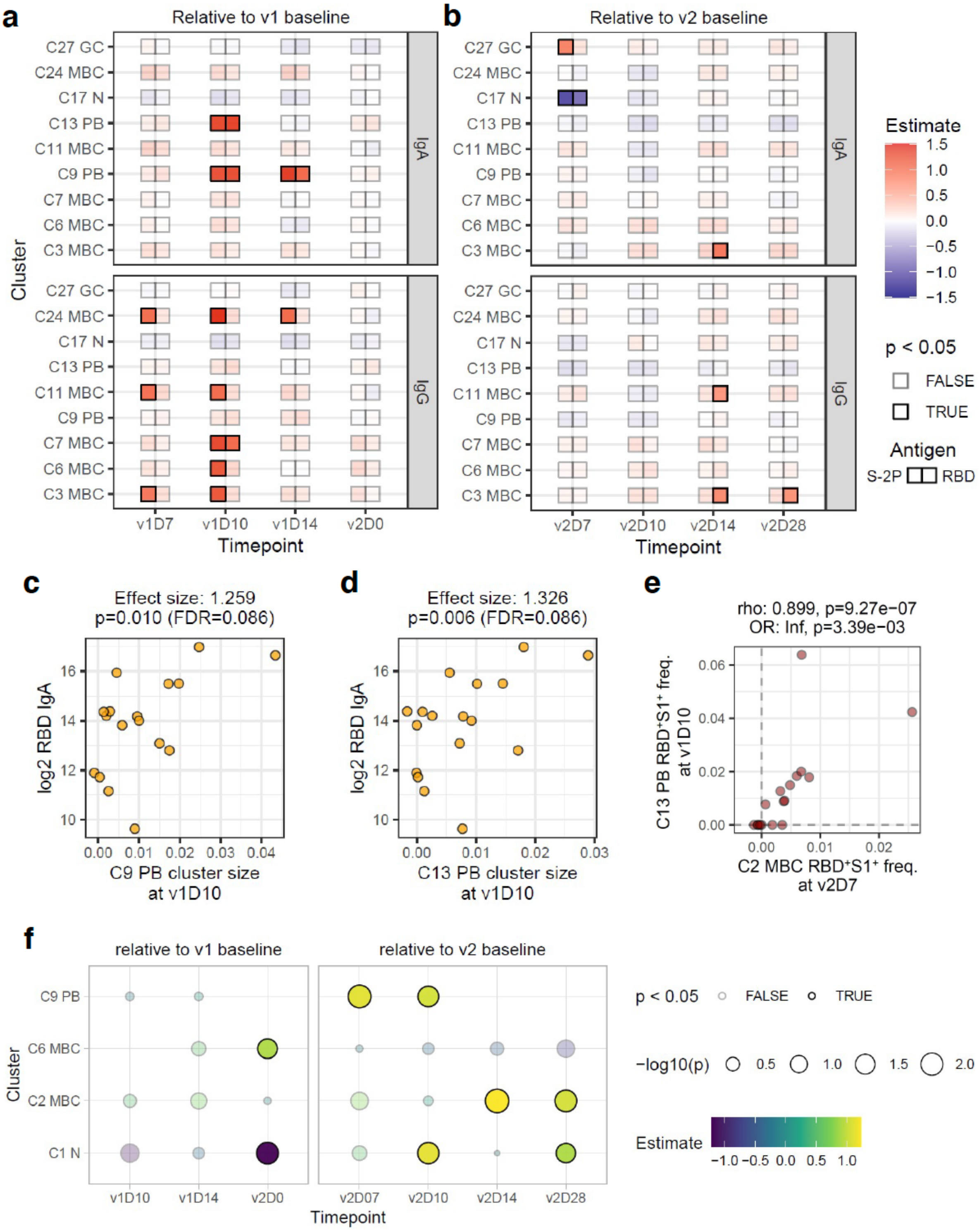
a
Relative to v1 baseline
b
Relative to v2 baseline
C27 GC
C24 MBC
C17 N
C13 PB
C11 MBC
C9 PB
C7 MBC
C6 MBC
C3 MBC
IgA
IgG
Cluster
v1D7
v1D10
v1D14
v2D0
v2D7
v2D10
v2D14
v2D28
Timepoint
Estimate
1.5
1.0
0.5
0.0
−0.5
−1.0
−1.5
p < 0.05
FALSE
TRUE
Antigen
S-2P
RBD
c
Effect size: 1.259
p=0.010 (FDR=0.086)
log2 RBD IgA
C9 PB cluster size at v1D10
d
Effect size: 1.326
p=0.006 (FDR=0.086)
log2 RBD IgA
C13 PB cluster size at v1D10
e
rho: 0.899, p=9.27e−07
OR: Inf, p=3.39e−03
C13 PB RBD⁺S1⁺ freq. at v1D10
C2 MBC RBD⁺S1⁺ freq. at v2D7
f
relative to v1 baseline
relative to v2 baseline
C9 PB
C6 MBC
C2 MBC
C1 N
Cluster
v1D10
v1D14
v2D0
v2D07
v2D10
v2D14
v2D28
Timepoint
p < 0.05
FALSE
TRUE
−log10(p)
0.5
1.0
1.5
2.0
Estimate
−1.0 −0.5 0.0 0.5 1.0

27

**Extended Data Fig. 3. Additional associations between cluster frequencies and endpoint SARS-CoV-2 antibody titers. a, b,** Similar to Fig. 4a and b, but for associations with antigen non-specific cells. **c, d,** Similar to Fig. 4c, but showing association between endpoint RBD IgA titers and antigen non-specific cells, i.e., cluster size as a fraction of total $CD19^+$ cell counts, within C9 **(c)**, and C13 **(d)**, on v1D10. **e,** Correlation between $RBD^+S1^+$ cell frequencies within C2 on v2D7 and C13 on v1D10. Spearman's correlation coefficients (rho) and unadjusted P values shown. **f,** Similar to Fig. 4f, but showing association between the first principal component (PC1) of endpoint SARS-CoV-2 antibody titers and spike-specific $RBD^+S1^+$ (double positive) cell frequencies within each cell cluster. D, day; FDR, false discovery rate; OR, odds ratio; RBD, receptor binding domain; S1, spike subunit 1; v, vaccine dose.

### Extended Table 1: Participant information and visits

| Participant | Age Range at Vaccination | Gender at Birth | Vaccine Dose 1# | | | | Vaccine Dose 2# | | | | | | Days Between Doses |
|---|---|---|---|---|---|---|---|---|---|---|---|---|---|
| | | | D0 | D7 | D10 | D14 | D0 | D5 | D7 | D10 | D14 | D28 | |
| VAC-001 | 51-55 | M | 1 | 7 | 9 | 14 | 0 | | 6 | 9 | 13 | 28 | 28 |
| VAC-002 | 56-60 | F | 0 | 7 | 11 | 14 | 0 | | 7 | 9 | 14 | 28 | 27 |
| VAC-003 | 61-65 | F | 0 | 6 | 9 | 14 | 0 | | 6 | 9 | 15 | 28 | 28 |
| VAC-004 | 51-55 | F | 0 | 7 | 9 | 15 | -1 | | 6 | 9 | 13 | 28 | 28 |
| VAC-005 | 61-65 | F | 0 | 7 | 11 | 14 | 0 | | 6 | 11 | 14 | 33 | 28 |
| VAC-611 | 51-55 | F | -21 | 7 | 10 | 14 | 0 | | 7 | 9 | 14 | 28 | 29 |
| VAC-613 | 56-60 | F | 2 | 7 | 10 | 15 | 0 | | 7 | 10 | 13 | 29 | 28 |
| VAC-638 | 56-60 | F | 0 | 7 | 10 | 14 | 0 | 5 | 7 | | 14 | 31 | 31 |
| VAC-662 | 51-55 | M | 0 | | | 15 | 0 | 5 | 7 | 9 | 14 | 28 | 28 |
| VAC-676 | 51-55 | M | 0 | 8 | | 15 | -1 | | 6 | 10 | 14 | 31 | 29 |
| VAC-683 | 61-65 | M | 0 | 7 | 10 | 15 | 0 | | 7 | 9 | 14 | 28 | 30 |
| VAC-685 | 41-45 | M | -1 | 7 | 10 | 14 | 0 | | 7 | 10 | 14 | 28 | 28 |
| VAC-687 | 51-55 | F | 0 | 8 | 11 | 14 | 0 | | 7 | 11 | 14 | 28 | 28 |
| VAC-713* | 41-45 | M | 1 | 6 | | 14 | | | | | | | |
| VAC-715 | 41-45 | F | 0 | 7 | | | 0 | | 7 | 10 | 14 | 28 | 25 |
| VAC-716 | 41-45 | F | 0 | 7 | 12 | 14 | 0 | | 7 | 11 | 14 | 28 | 29 |
| VAC-717 | 56-60 | F | | 7 | 11 | 14 | 0 | 5 | 7 | 9 | | 28 | 34 |
| VAC-718 | 36-40 | F | -1 | 7 | 11 | | 0 | | 7 | 11 | 13 | 31 | 28 |
| VAC-719 | 56-60 | M | | 7 | 11 | 14 | 0 | | 7 | 11 | 14 | 28 | 28 |
| VAC-720 | 36-40 | M | 0 | 7 | 10 | 14 | 0 | | 6 | 11 | 14 | 28 | 27 |
| VAC-721 | 61-65 | M | | 7 | 10 | 15 | -1 | | 6 | 10 | 12 | 26 | 26 |
| **Median†/N** | **55†** | **9M/11F** | **18** | **20** | **17** | **19** | **20** | **3** | **20** | **19** | **19** | **28** | **28†** |

*Lost to follow-up after contracting COVID-19 after dose 1
#Numbers indicate actual date of visit relative to vaccination day

**Extended Data Table 2: 17-color flow cytometry panel**

| Reagent | Clone | Source | Catalogue Number | Dilution |
|---|---|---|---|---|
| Mouse anti-human CD11c BUV395 | B-ly6 | BD Biosciences | 563787 | 1:100 |
| Mouse anti-human CD138 BUV737 | MI15 | BD Biosciences | 612834 | 1:200 |
| Mouse anti-human CD45 BUV805 | HI30 | BD Biosciences | 612891 | 1:100 |
| Mouse anti-human CD38 BV421 | HIT2 | BD Biosciences | 562444 | 1:100 |
| Mouse anti-human CD19 BV650 | SJ25-C1 | BD Biosciences | 563226 | 1:100 |
| Mouse anti-human CD10 BV510 | HI10a | BD Biosciences | 563032 | 1:100 |
| Mouse anti-human CD3 BV570 | UCHT1 | Biolegend | 300436 | 1:100 |
| Mouse anti-human IgD BV605 | IA6–2 | Biolegend | 348232 | 1:100 |
| Mouse anti-human IgM BV711 | MHM-88 | Biolegend | 314540 | 1:100 |
| Mouse anti-human CD14 BV750 | 63D3 | Biolegend | 367136 | 1:100 |
| Mouse anti-human CD27 BV785 | O323 | Biolegend | 302832 | 1:100 |
| Mouse anti-human CD21 FITC | BU32 | Biolegend | 354910 | 1:100 |
| Mouse anti-human IgG PE-Cy7 | G18–145 | BD Biosciences | 561298 | 1:100 |
| Mouse anti-human CD20 APC-H7 | 2H7 | BD Biosciences | 560734 | 1:100 |
| SARS-CoV-2 S protein S1 | | Biolegend | 793806 | |
| SARS-CoV-2 S protein RBD | | In-house | | |

### Extended Data Table 3: Detailed cluster annotations and designations

| Cluster | Population | Ig isotype | Defining markers | Designations/other names | References |
|---|---|---|---|---|---|
| 28 | I/T | IgM>D | $CD10^{+}CD38^{++}CD27^{-}CD138^{+}$ | | |
| 16 | N-GC | IgD>M | $CD10^{+}CD38^{+}CD27^{lo}$ | | |
| 27 | GC | IgM>D | $CD10^{+}CD38^{++}CD27^{+}CD138^{lo}$ | GC founder | |
| 23 | MBC | IgD>M | $CD27^{lo}CD38^{-}CD20^{++}CD21^{lo}CD11c^{lo}$ | Nonconventional MBC/Atypical/TLM | 37,38 |
| 29 | MBC | IgM>D | $CD27^{+}CD38^{-}CD20^{++}CD21^{lo}CD11c^{lo}$ | Nonconventional MBC/Atypical/AM/ABC | 24,37,38 |
| 30 | MBC | IgM>D | $CD27^{+}CD38^{-}CD138^{+}$ | Conventional MBC | |
| 21 | N | IgD=M | $CD38^{-}$ | | |
| 19 | MBC | IgM>D | $CD27^{+}CD38^{-}$ | Conventional MBC | |
| 25 | MBC | IgM>D | $CD27^{+}CD38^{+}$ | Conventional MBC | |
| 22 | MBC | $IgD^{+}IgM^{-}$ | $CD27^{+}CD38^{-}$ | Conventional MBC/IgD only MBC | |
| 24 | MBC | IgD>M | $CD27^{lo}CD38^{-}CD20^{++}CD21^{lo}CD11c^{+}$ | Nonconventional MBC/Atypical/TLM | 37,38 |
| 15 | MBC | IgD>M | $CD27^{lo}CD38^{+}CD11c^{+}$ | Nonconventional MBC | |
| 20 | N-MBC | IgD>M; IgG | $CD27^{lo}CD38^{+}$ | Likely bound IgG | |
| 1 | N | IgD>M | $CD38^{+}$ | | |
| 17 | N | $IgD^{+}IgM^{-}$ | $CD38^{+}$ | | |
| 12 | pPB | IgA | $CD27^{lo}CD38^{++}CD20^{-}CD21^{lo}$ | | |
| 13 | PB | IgA | $CD27^{+}CD38^{+++}CD20^{-}CD21^{lo}$ | | |
| 9 | PB | IgG | $CD27^{+}CD38^{+++}CD20^{-}CD21^{lo}$ | | |
| 8 | NB | Multiple Ig | $CD10^{+}CD14^{lo}$ | Granulocyte | |
| 5 | MBC | IgG | $CD27^{+}CD38^{-}CD20^{++}CD21^{lo}CD11c^{lo}$ | Nonconventional MBC/Atypical/AM/ABC | 24,37,38 |
| 3 | MBC | IgG | $CD27^{-}CD38^{-}CD20^{++}CD21^{lo}CD11c^{+}$ | Nonconventional MBC/Atypical/TLM/DN2 | 37-39 |
| 4 | MBC | IgG | $CD27^{+}CD38^{-}$ | Conventional MBC | |
| 2 | MBC | IgG | $CD27^{+}CD38^{+}$ | Conventional MBC | |
| 7 | MBC | IgG | $CD27^{-}CD38^{-}$ | Nonconventional MBC | |
| 6 | MBC | IgG | $CD27^{-}CD38^{+}$ | Nonconventional MBC | |
| 11 | MBC | IgA | $CD27^{lo}CD38^{-}CD20^{++}CD21^{-}CD11c^{+}$ | Nonconventional MBC/Atypical/TLM/DN2 | 37-39 |
| 14 | MBC | IgA | $CD27^{+}CD38^{+}$ | Conventional MBC | |
| 26 | MBC | $IgM^{+}IgD^{-}$ | $CD27^{+}CD38^{+}$ | Conventional MBC/IgM only memory | |
| 18 | N | Low multiple Ig | $CD38^{+}$ | | |
| 10 | MBC | Low multiple Ig | $CD27^{+}CD38^{lo}$ | Conventional MBC | |

ABC, activated B cell; AM, activated memory; DN2 double negative 2; GC, germinal center; I/T immature/transitional; MBC, memory B cell; N, naïve; NB, not B cell; PB, plasmablast; pPB, pre-PB; TLM, tissue-like memory

**Extended Data Table 4: COVID-19 patient information**

| Patient | Age Range at enrollment | Gender at birth | Disease day* of sample |
|---|---|---|---|
| 4001 | 36-40 | M | 20 |
| 4004 | 21-25 | M | 34 |
| 4005 | 26-30 | F | 35 |
| 4007 | 61-65 | M | 19 |
| 4008 | 51-55 | F | 27 |
| 4009 | 56-60 | M | 24 |
| 4301 | 66-70 | M | 39 |
| 4302 | 41-45 | F | 29 |
| 4303 | 36-40 | M | 23 |
| 002 | 71-75 | M | 73 |
| 003 | 36-40 | M | 35 |
| 004 | 26-30 | M | 36 |
| 005 | 26-30 | F | 38 |
| 007 | 81-85 | M | 32 |
| 008 | 56-60 | F | 35 |
| 010 | 51-55 | M | 70 |
| 011 | 46-50 | M | 17 |
| 012 | 71-75 | M | 34 |
| 013 | 56-60 | M | 37 |
| 014 | 46-50 | F | 18 |
| 015 | 41-45 | M | 15 |
| **Median[†]/N** | **48[†]** | **15M/6F** | **34[†]** |

*At peak antibody response or latest timepoint available

**Extended Data Table 5: Additional reagents for flow cytometric analyses**

| Reagent | Clone | Source | Catalogue Number | Dilution |
|---|---|---|---|---|
| Mouse anti-human CD3 BV510 | OKT3 | Biolegend | 317332 | 1:100 |
| Mouse anti-human CD27 BV421 | O323 | Biolegend | 302824 | 1:100 |
| Mouse anti-human IgD PE-Cy7 | IA6-2 | Biolegend | 348210 | 1:100 |
| Mouse anti-human IgM APC | MHM-88 | Biolegend | 314510 | 1:200 |
| Mouse anti-human IgG PE | G18–145 | BD Biosciences | 555787 | 1:40 |
| Mouse anti-human IgA FITC | IS11-8E10 | Miltenyi Biotec | 130-113-475 | 1:200 |
| Mouse anti-human CD19 PerCP-Cy5.5 | SJ25-C1 | ThermoFisher | 45-0198-42 | 1:100 |
| Streptavidin-PE | | ThermoFisher | S21388 | |
| Streptavidin-APC | | ThermoFisher | S32362 | |
| Streptavidin-Alexa Fluor 488 | | ThermoFisher | S32354 | |
| SARS-CoV-2 S2 super stable trimer | | AcroBiosystems | SPN-C82E9 | |
